# Self-perceived cognition and psychological well-being in patients with chronic kidney disease undergoing maintenance haemodialysis

**DOI:** 10.64898/2026.10.02.26364564

**Authors:** Alexandra-Elena Marin, Rosa Redolat, Alicia García-Testal, Eva Segura-Ortí, Patricia Mesa-Gresa

**Affiliations:** Department of Psychobiology, Faculty of Psychology and Logopedics, Universitat de Valencia, Valencia, Spain; Hospital at Home, Hospital Clínico Universitario de Valencia, Valencia, Spain; Department of Nephrology, Hospital de Manises, Manises, Spain; Universidad Cardenal-Herrera-CEU, CEU Universities, Physiotherapy, Alfara del Patriarca, Spain

**Author notes:** **Corresponding author:** Patricia Mesa-Gresa, PhD, Full Professor, Department of Psychobiology, Faculty of Psychology and Logopedics, Universitat de Valencia, Valencia, Spain. **Email addresses:** Alexandra-Elena MARIN, Rosa REDOLAT, Alicia GARCÍA-TESTAL, Eva SEGURA-ORTÍ.

**Keywords:** chronic kidney disease, haemodialysis, self-perceived cognition, psychological well-being, patient-reported outcomes, biomarkers

## Abstract

Cognitive and psychological difficulties are frequently reported by patients undergoing haemodialysis, yet these experiences are rarely explored within routine clinical assessments. The aim of this study was to examine self-perceived cognition and psychological well-being as patient-reported outcomes in adults with chronic kidney disease undergoing maintenance haemodialysis, and to explore the relationships between these outcomes and sociodemographic, clinical, and biochemical variables. Adults receiving maintenance haemodialysis were recruited from a hospital-based dialysis unit as part of the GoodRENal project, which focuses on developing educational approaches for people on haemodialysis. Participants completed self-report measures assessing perceived cognitive functioning and psychological well-being. Sociodemographic, clinical, and biochemical data routinely collected in dialysis care were obtained from medical records. Associations were examined between self-perceived cognition, psychological well-being, and the available data. Seventy-eight patients took part in the study. Perceived cognitive difficulties were more closely related to indicators of kidney disease severity and metabolic imbalance, whereas psychological well-being showed stronger associations with social and emotional factors. The patients’ subjective experiences could not be fully explained by objective clinical indicators alone, suggesting a gap between physiological measures and patient-reported outcomes. Self-perceived cognition and psychological well-being provide clinically relevant information that could complement routine clinical indicators in haemodialysis care.

## 1. INTRODUCTION

Chronic kidney disease (CKD) is a condition characterized by the alteration of kidney structure and function lasting more than three months, that affects more than 10% of the world’s population with a higher prevalence among older individuals, women and racial minorities (García et al., 2022). Current estimates suggest that over 800 million people have been diagnosed with CKD, and projections indicate that by 2040 this disease will rank fifth in terms of global years of life lost. These projections are significant because the number of people with CKD may not accurately reflect the true prevalence of the disease. The fact that CKD is usually asymptomatic until the disease is advanced, makes it difficult to calculate the true burden and its exact prevalence (Jha et al., 2013). Recent evidence indicates that in certain countries such as Spain, Australia or Brazil, the numbers of people with undiagnosed CKD stage 3 ranges from 85 to 97% (Pecoits-Filho et al., 2023). Furthermore, CKD is linked with increased mortality and morbidity, representing a significant burden particularly for patients living in low-and middle-income countries that lack the resources to address its detrimental consequences (Kalantar-Zadeh, 2021).

A diagnosis of CKD is made when a patient shows signs of kidney damage such as structural abnormalities, haematuria or a history of kidney transplantation, or when their glomerular filtration rate (GFR) falls below 60 ml/min/1.73 m2 for more than three months. As a result, the five different stages of CKD are determined based on the basis of GFR, with advanced chronic kidney disease (ACKD) being recognized as occurring between stages 4 and 5, and CKD Stage 5 Dialysis-Dependent (CKD 5D) being the last stage of CKD. Once this point is reached, the disease becomes irreversible, and the primary treatment options include kidney transplant or the replacement of kidney function through peritoneal dialysis (PD) or hemodialysis (HD) (Tangri et al., 2023).

Patients diagnosed with CKD often present comorbidities, namely hypertension, diabetes mellitus and cerebrovascular diseases. These diseases, along with metabolic factors, contribute to the development of cognitive impairment (CI) within patients with CKD (Sudrajat et al., 2022). As CKD progresses, CI gradually emerges and becomes highly prevalent in advanced stages of the disease, affecting nearly two-thirds of individuals with CKD 5D. Moreover, CKD 5D has been identified as the third most important risk factor for mild cognitive impairment (MCI) and dementia, after stroke and chronic use of anxiolytics, putting it ahead of other factors such as genetics (Lipnicki et al., 2017).

The cognitive processes most affected in CKD include attention, orientation, memory, concentration, language, and processing speed (Mesa-Gresa et al., 2024). These cognitive skills support an individual’s ability to organise, monitor, and execute goals (Drew et al., 2019). As a result, CI is closely associated with reduced engagement in healthcare and poorer disease self-management (Mc-Adams-DeMarco et al., 2020). The extent of cognitive decline varies by dialysis modality, with patients undergoing HD showing more severe impairment and higher susceptibility to dementia than those receiving peritoneal dialysis (Tian et al., 2019). Up to 80% of individuals undergoing haemodialysis report cognitive impairment, with advanced age, cardiovascular disease, and metabolic disorders as common risk factors (Dasgupta et al., 2018). However, more recent findings suggest a slightly lower but still substantial prevalence of CI of around 70%, reinforcing the significance of this issue within the CKD 5D population (Andrews et al., 2025).

Despite the high prevalence of dementia-like conditions in patients with CKD 5D, these conditions often go undiagnosed due to a variety of factors, including medical focus, lack of awareness and overlapping symptoms (Coyne et al., 2023). For a better neurological evaluation, it is important not only to use standardized screening tests but also to take into account the self-reports regarding patients’ daily functioning and cognitive-related aspects, such as memory, attention or concentration (Chan et al., 2022; Chan et al., 2024).

As noted above, CKD 5D and its associated pathologies, including CI, impose a significant burden on those affected by this disease. This burden is further compounded by an increased susceptibility to mood disorders, such as anxiety and depression (Huang et al., 2021; Marin et al., 2023). In fact, depression in people undergoing dialysis treatment or living with kidney transplant increases the risk of hospitalization and death by about 50% (Cogley et al., 2022). Although depression is significantly associated with CI (Feng et al., 2022; Joseph et al., 2019), there is a paucity of data pertaining to the different domains of cognitive impairment, anxiety and depression, and their association with clinical parameters in individuals undergoing HD (Golenia et al., 2023).

The relationship between CKD 5D, cognitive decline, and mood disorders is complex, and recent research has focused on biomarkers associated with cognitive impairment, anxiety, and depression in this population. These biomarkers range from indicators of neurodegeneration and inflammation to kidney disease–specific markers. Chi et al. (2023) examined associations between kidney disease, blood markers, and the risk of dementia or cognitive decline, identifying elevated serum creatinine and urinary albumin–creatinine ratio as risk markers.

The literature on the effect of albuminuria on dementia risk in renal impairment is also extensive, with many studies indicating that high albumin levels can be associated with an increased likelihood of developing cognitive decline or dementia (Drew et al., 2019).

The role of uric acid in cognitive decline is complex and influenced by several factors, including population characteristics and gender. While some studies suggest a protective effect (Ye et al., 2016), others associate both low and high levels with increased cognitive risk. In CKD, evidence increasingly links elevated uric acid to poorer cognitive function (Liabeuf et al., 2021; Sudrajat et al., 2022; Tian et al., 2019).

Cognitive impairment in CKD is also associated with higher levels of inflammation, with C-reactive protein being associated with poorer cognitive status (Kaltsatou, 2016); higher levels of Hemoglobin A1c (HbA1c), are associated with poorer memory and executive function (Seidel et al., 2014). With regard to phosphorus, this essential nutrient is crucial for energy production and appears to play a critical role in maintaining brain health (Bird & Eskin, 2021). A review by Rroji et al. (2022) examines the pathophysiological effects of phosphate toxicity on the nervous system and suggests that elevated phosphate levels may play a role in CI, particularly in people with CKD.

A recent review of the patterns of CI in patients undergoing HD and the associated factors found a high prevalence of CI, depression and anxiety symptoms in this patient cohort (Golenia et al., 2023). The researchers also examined the relationship between these variables and clinical factors. They found that patients with anxiety symptoms tended to be younger and have a lower educational level than those who experience lower levels of anxiety. These patients also exhibited physiological parameters such as higher levels of hemoglobin, parathyroid hormone (PTH) and endogenous creatinine (Golenia et al., 2023). These findings are consistent with previous studies showing a positive association between elevated creatinine levels and increased stress and psychological distress (Guerra et al., 2021). Conversely, a systematic review by Huang et al. (2021) and a study by Bossola et al. (2010) reported that lower PTH levels were associated with increased anxiety symptoms. The latter study also found that depressive symptoms correlated positively with age, and negatively with creatinine and serum albumin levels.

As reviewed in the previous sections, different studies have evaluated the objective cognitive and psychological well-being of patients with CKD 5D, as well as its relationship with different clinical parameters. However, the self-perceived state of these patients has not been extensively evaluated. The current study aims to assess patients’ subjective perceptions of their cognitive functioning and psychological well-being, and to describe the characteristics of a specific sample of patients undergoing HD. In addition, we aim to examine the relationship between the patients’ self-perceived cognitive and psychological state and their sociodemographic, clinical and biochemical variables. Regarding cognition, we hypothesize that higher levels of uric acid (Sudrajat et al., 2022; Tian et al., 2019), albumin (Drew et al., 2019), creatinine (Chi et al., 2023), hemoglobin A1c (Seidel et al., 2014) and phosphorus (Rroji et al., 2022) will be linked to a poorer self-perceived cognitive state. Regarding psychological well-being, we expect that higher levels of creatinine, hemoglobin and PTH (Golenia et al., 2023), and lower levels of serum albumin (Bossola et al., 2010) will be associated with a lower self-perceived psychological well-being in patients on HD. Lastly, we hypothesize that sociodemographic and clinical variables will also play a role in shaping patients’ perceptions of their own cognitive and psychological state.

## 2. METHODS

### 2.1. STUDY DESIGN AND PARTICIPANTS

This study adopts a cross-sectional observational approach in accordance with the design proposed by Strengthening the Reporting of Observational Studies in Epidemiology (STROBE Statement) (Von Elm et al., 2008), which entails a checklist of essential elements to be included in reports of observational studies.

A group of 78 patients undergoing HD (age 68.3 ± 14.0 years; women 50%) were selected from the Hospital de Manises in Valencia, Spain. The inclusion criteria were as follows: 1) individuals with CKD who had undergone regular HD treatment for a minimum of 3 months; 2) aged 18 years or older; 3) individuals who received at least two HD sessions per week; 4) individuals free from unstable conditions such as delirium or dementia, or other significant physical or psychological disabilities that could invalidate informed consent or their responses. The participants underwent HD sessions two to three times per week, with each session lasting approximately 4 hours.

### 2.2. STUDY SETTING

The medical staff of Hospital de Manises identified suitable patients who were then enrolled in the study during the dialysis therapy, taking into account inclusion criteria. After being fully informed about the study, written informed consent was obtained from and signed by all participants prior to their participation. The exploratory questionnaires were administered in the dialysis unit by trained healthcare professionals.

### 2.3. DATA COLLECTION

Sociodemographic variables (age, gender, relationship status, number of children, actively working, tobacco use, alcohol use, psychotropic drug use, body mass index) and clinical data (etiology of CKD, current vascular access, hypertension, cardiovascular disease, diabetes mellitus, active neoplastic disease) were collected from the patients’ medical records. Blood samples were obtained before dialysis during one of the patients’ regular mid-week sessions, as part of the routine monthly blood tests typically performed in patients undergoing HD. Laboratory data regarding creatinine, albumin, uric acid, phosphorus, PTH, C-reactive protein, hemoglobin, hemoglobin A1c, calcium, sodium, potassium, cholesterol low-density lipoprotein (LDL), cholesterol high-density lipoprotein (HDL), cholesterol total, triglycerides, leukocytes, glucose and thyroid stimulating hormone (TSH) were recorded. The values for laboratory variables were calculated as the means of different measurements for each variable taken over the preceding 6 months.

Two exploratory questionnaires designed ad-hoc by experienced health professionals working with patients on HD were used for this study (Mesa-Gresa et al., 2023). The aim of these questionnaires was to assess both the patients’ subjective perception of their psychological well-being and the patients’ self-perceived cognitive state over the past month. The questionnaire evaluating cognitive function assessed self-perceived executive functions, memory and attention, among others. Furthermore, psychological well-being included aspects such as mood, anxiety, stress response, overload, sleep quality and motivation. Each questionnaire consisted of 10 questions, formulated using a 5-point Likert scale with the following options: 1 (never), 2 (now and then), 3 (half of the time), 4 (more than half of the time), and 5 (almost all the time). Additionally, an open-ended question was included to assess barriers, difficulties and needs. The questions were aimed at detecting cognitive impairment or emotional distress. To ensure that total higher scores indicated a better condition, certain items were reverse scored. Specifically, in the cognitive section, items 1–6 and 9 were reversed, while in the psychological well-being section, items 3–5 and 8 were adjusted accordingly. For the analysis of the data obtained from the questionnaires, responses with scores of 4 and 5 points, indicating a positive level of agreement, were grouped together to facilitate interpretation.

To minimize information bias, validated self-report instruments were used, and clinical and biochemical data were extracted from routinely collected medical records.

### 2.4. DATA ANALYSIS

SPSS statistical software version 27.0, (IBM Corp, Armonk, NY, USA) was used to analyze the data. Descriptive statistics are presented as numbers and percentages for categorical variables, and continuous data are presented as mean ± standard deviation. One-way ANOVAs were performed to examine the effect of independent variables such as sociodemographic variables (group of age, gender, relationship status, etc. See Table 1), clinical variables (etiology of CKD, hypertension, diabetes mellitus, etc. See Table 1) and biomarkers (creatinine, uric acid, phosphorus, etc. See Table 2) on self-perceived cognitive state and psychological state, independently. Pearson correlations were used to analyze correlations between 1) sociodemographic variables and total questionnaire scores; 2) sociodemographic variables and laboratory data; 3) laboratory data and total questionnaire scores; 4) different laboratory variables; and 5) total cognition score and total psychological well-being score. Standard multiple regression was used to assess the independent influences of creatinine, phosphorus and uric acid on self-perceived cognitive state. No analyses were carried out to examine potential subgroup or interaction effects.

**Table 1.**
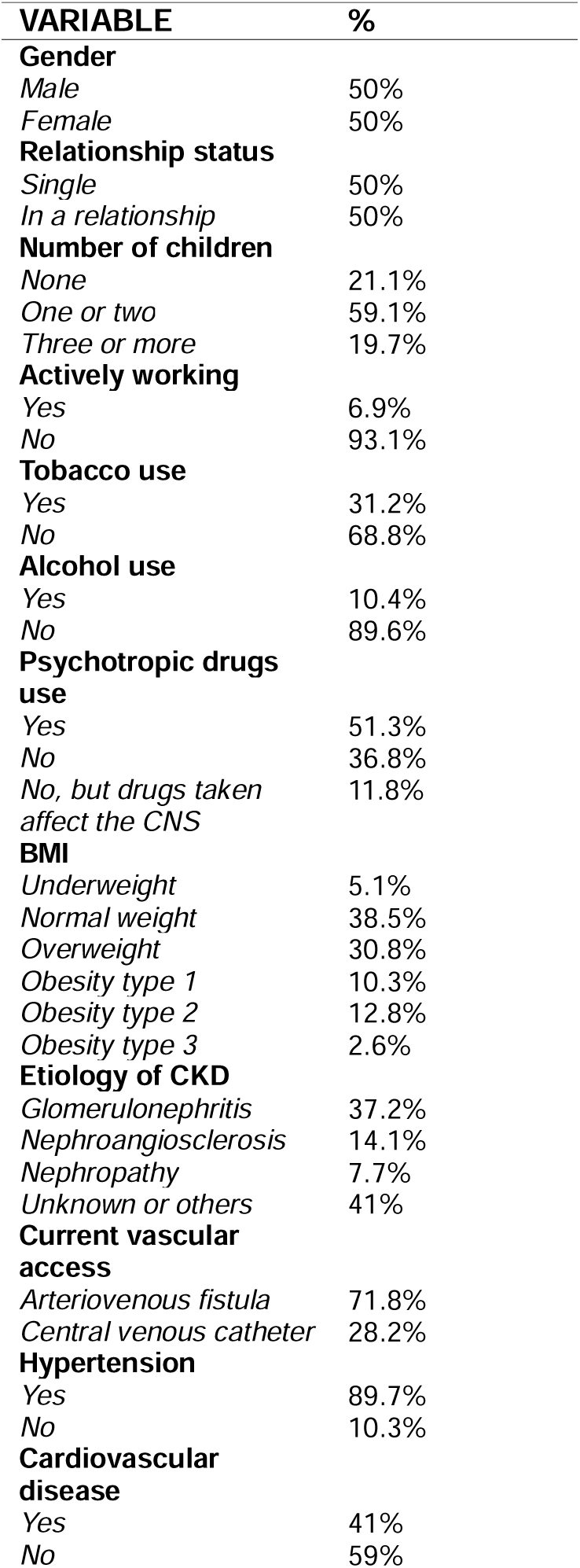
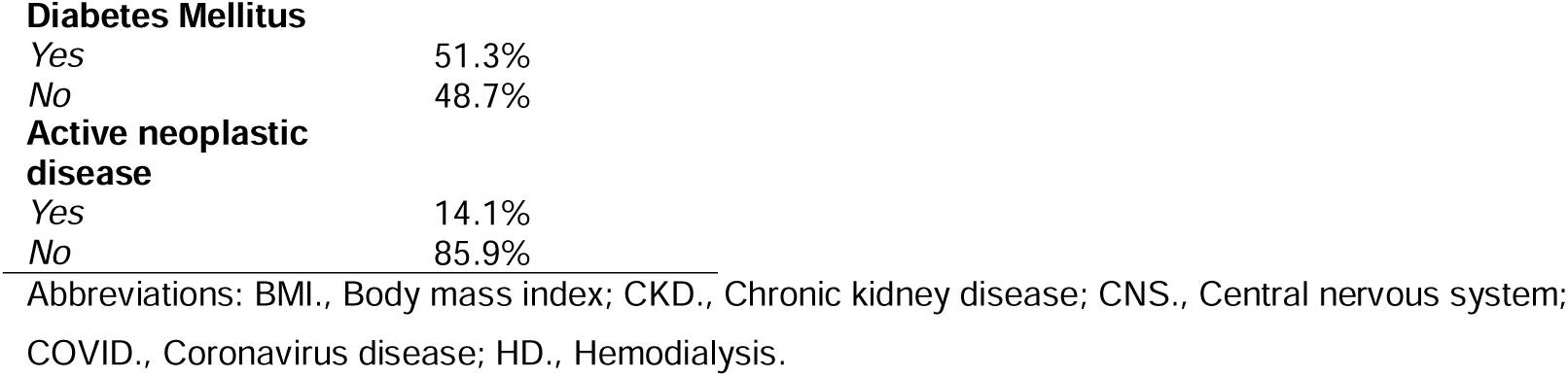
Sociodemographic and clinical characteristics of patients with chronic renal failure undergoing haemodialysis.

**Table 2.** Laboratory characteristics of CKD patients with chronic renal failure undergoing haemodialysis.

| **VARIABLE** | **MEAN** | **SD** | **MIN.** | **MAX.** |
| --- | --- | --- | --- | --- |
| Creatinine, mg/dL | 6.94 | 1.87 | 3.1 | 12.4 |
| Albumin, g/dL | 3.86 | .31 | 2.8 | 4.5 |
| Uric Acid, mg/dL | 5.64 | 1.35 | 3.2 | 9.7 |
| Phosphorus, mg/dL | 4.43 | 8.0 | 1.9 | 1.11 |
| PTH (Corrected), pg/mL | 248.04 | 97.36 | 110.0 | 668.2 |
| C-Reactive Protein, mg/L | 11.70 | 28.13 | .0 | 205.9 |
| Hemoglobin, g/dL | 11.54 | .60 | 10.1 | 13.4 |
| Hemoglobin A1c (%) | 5.83 | 1.09 | 4.4 | 9.3 |
| Calcium, mg/dL | 9.00 | .46 | 7.9 | 10.1 |
| Sodium, mmol/L | 138.76 | 2.47 | 129.8 | 144.8 |
| Potassium, mmol/L | 5.03 | .57 | 3.0 | 6.2 |
| Cholesterol LDL, mg/dL | 64.64 | 28.08 | 12 | 170 |
| Cholesterol HDL, mg/dL | 41.31 | 11.09 | 19 | 68 |
| Cholesterol Total, mg/dL | 132.49 | 30.39 | 67 | 232 |
| Triglycerides, mg/dL | 139.01 | 57.48 | 47 | 288 |
| Leukocytes, x 10<sup>3</sup>/μL | 7.37 | 2.68 | 2.7 | 18.9 |
| Glucose, mg/dL | 146.60 | 68.01 | 62 | 395 |
| TSH, mU/L | 2.01 | 1.39 | .010 | 9.33 |

Abbreviations: CKD., Chronic kidney disease; dL., Deciliter; g., Gram; HD., Hemodialysis; HDL., High-density lipoprotein; LDL., Low-density lipoprotein; L., Liter; Max., Maximum; mg., Milligram; Min., Minimum; mmol., Millimole; mU., Mega unit; pg., Picogram; PTH., Parathyroid hormone; SD., Standard deviation; TSH., Thyroid Stimulating Hormone.

For all the statistical tests, *p* < 0.05 was considered as statistically significant.

### 2.5. ETHICAL CONSIDERATIONS

The study adhered to the principles outlined in the Declaration of Helsinki and was approved by the Ethics Committee of the University of Valencia (Reference: 2023-PSILOG-2558999) and the ‘Hospital Universitario y Politécnico La Fe’ (Spain) (Registration number: 2020-064-1).

## 3. RESULTS

### 3.1. SOCIODEMOGRAPHIC AND CLINICAL CHARACTERISTICS OF THE SAMPLE

Seventy-eight adult patients were included in the study (age 68.3±14.0 years; women 50%). Autogenous arteriovenous fistulas was used in 56 patients, and central venous catheter was used in 22 patients. The HD vintage was 45.0 months, and the mean HD dialysis length was 218.9 minutes. The demographic and clinical characteristics of the patients (including characteristics associated with the dialysis treatment) are summarized in Table 1; and laboratory characteristics are summarized in Table 2. No substantial missing data were observed.

### 3.2. SELF-PERCEIVED COGNITIVE AND PSYCHOLOGICAL STATE

As can be seen in Table 3, patients’ general perceptions of their cognitive functioning were positive, with more than 50% of patients reporting a very good state for 8 out of the 10 items, assessed in the exploratory questionnaire about self-perceived cognitive state.

**Table 3.**
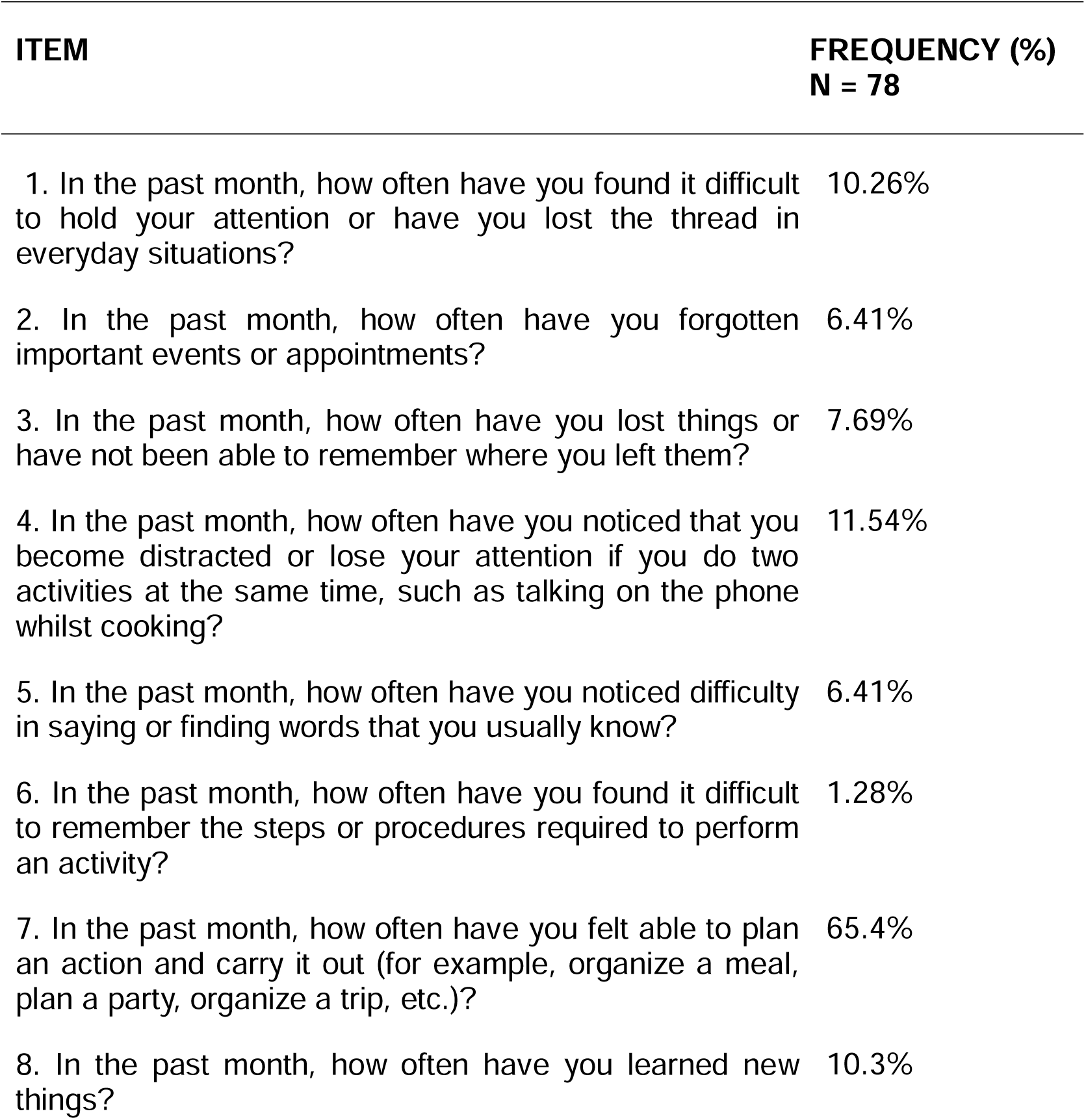
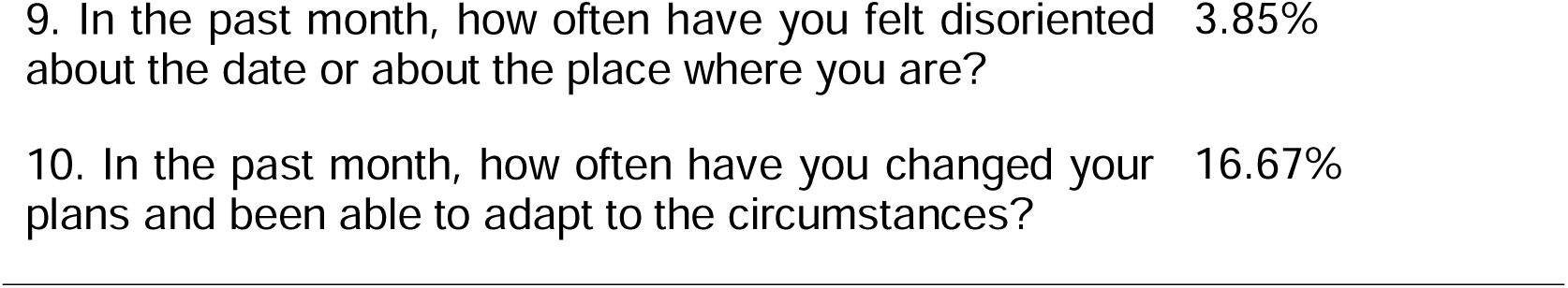
Exploratory questionnaire items regarding self-perceived cognitive state and number and percentage of people who agreed with the items “more than half of the time” and “most of the time” (4 and 5 scores). Note that items 1-6 and 9 are reversed.

Similarly to their self-perceived cognitive state, patients generally reported a relatively good psychological and emotional well-being, with over 50% of them reporting a good or a very good state for 5 out of the 10 items (Table 4).

**Table 4.**
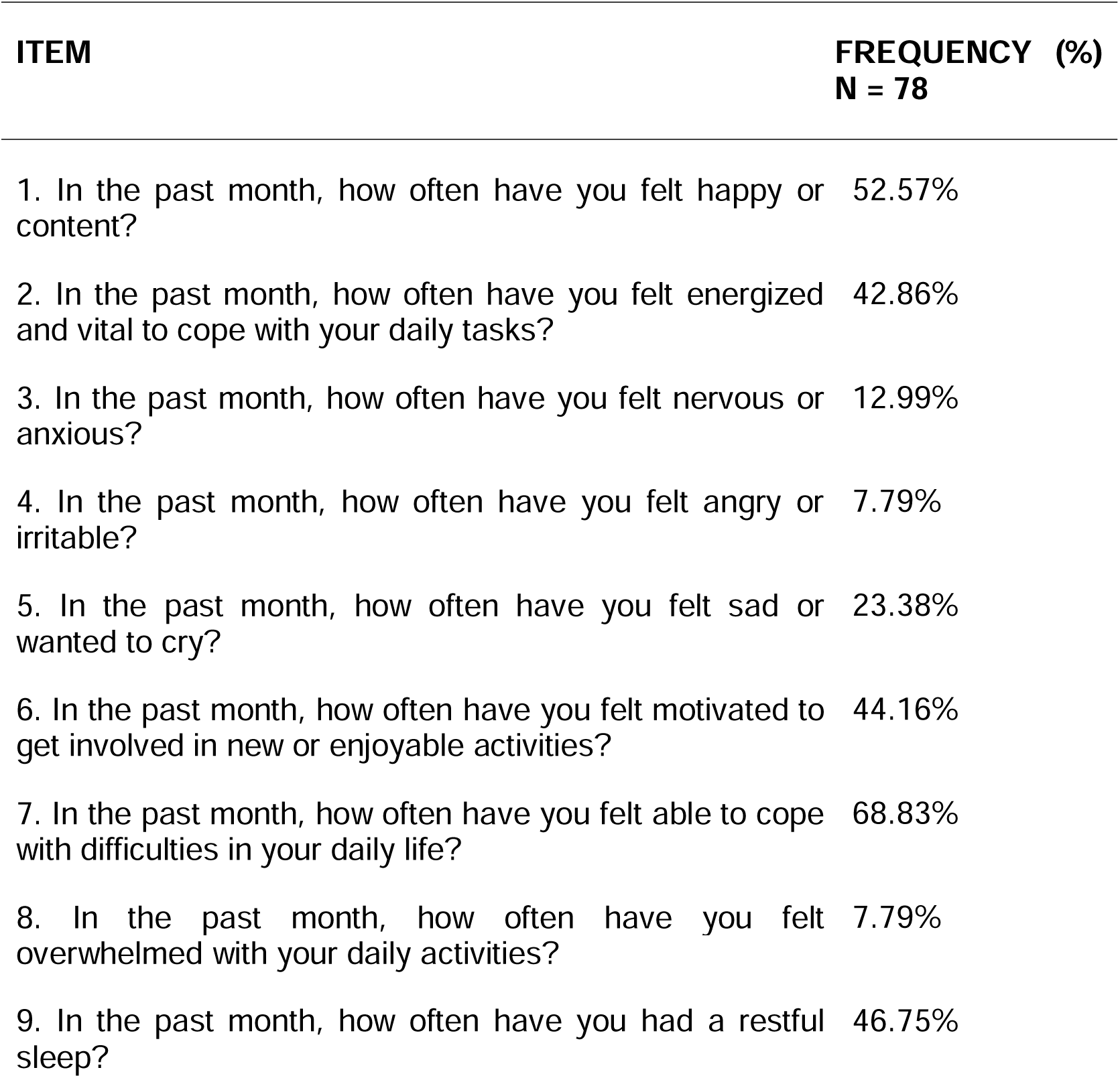
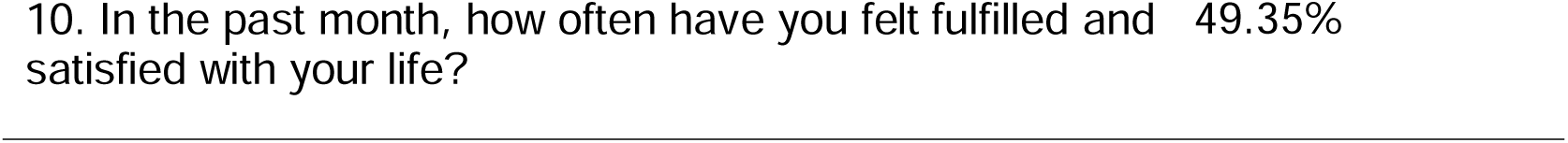
Items from the exploratory questionnaire regarding self-perceived psychological state and number and percentage of people who agreed with the item “more than half of the time” and “most of the time” (4 and 5 scores). Note that items 3-5 and 8 are reversed.

### 3.3. THE ASSOCIATION OF SELF-PERCEIVED COGNITIVE AND PSYCHOLOGICAL WELL-BEING WITH SOCIODEMOGRAPHIC, CLINICAL AND BIOCHEMICAL VARIABLES

#### 3.3.1. SELF-PERCEIVED COGNITIVE STATE AND SOCIODEMOGRAPHIC VARIABLES

A one-way between-groups analysis of variance was conducted to explore the impact of sociodemographic variables (age, gender, relationship status, number of children, actively working, tobacco use, alcohol use, psychotropic drug use, body max index) on self-perceived cognitive state. The results revealed a significant difference in the total cognition score [*F*(1, 70) = 6.208, *p* = .015], with a worse self-perceived cognitive status was observed in single patients (*M* = 37.50, *SD* = 5.23) compared to those in a relationship (*M* = 40.14, *SD* = 3.61). The effect size, calculated using eta squared, was .08. No other significant results were found for the remaining sociodemographic variables.

#### 3.3.2. SELF-PERCEIVED COGNITIVE STATE AND CLINICAL VARIABLES

A one-way between-groups analysis of variance was performed to examine the effect of clinical variables (etiology of CKD, current vascular access, hypertension, cardiovascular disease, diabetes mellitus, active neoplastic disease) on self-perceived cognitive state. The results revealed a significant difference in the total cognition score [*F*(1, 76) = 5.873, *p* = .020], with a worse self-perceived cognitive state in patients who did not have diabetes (*M* = 37.11, *SD* = 6.03) compared to those who did have it (*M* = 39.83, *SD* = 3.65). The effect size, calculated using eta squared, was .07. No other significant results were found for the remaining clinical variables.

#### 3.3.3. SELF-PERCEIVED COGNITIVE STATE AND BIOCHEMICAL VARIABLES

The relationship between the different biomarkers evaluated and self-perceived cognitive state was investigated using Pearson correlation coefficient. Significant correlations were obtained between some of these variables and the total score of the exploratory questionnaire. Particularly, there were significant negative relationships between self-perceived cognitive state and levels of creatinine (*r* = −.250; *p =* .027); phosphorus (*r* = −.276; *p* = .014); and uric acid (*r* = −306; *p* = .007). No other significant results were found.

Furthermore, we wanted to assess if these three variables together (creatinine, phosphorus and uric acid) could be used to predict the outcome of the questionnaire measuring self-perceived cognitive state. To do this, we used a standard multiple regression. Preliminary analyses were carried out to ensure no violation of the assumptions of normality, linearity, multicollinearity and homoscedasticity. Results revealed that the three variables explained a significant proportion (9.8%) of the variance in self-perceived cognitive state [*F*(3, 72) = 3.702, *p* = .015, R^2^ = .098]. Beta coefficients, standard errors, standardized betas and significance value for each predictor can be seen in Table 5. Uric acid made the strongest unique contribution to explaining the self-perceived cognitive state of the participants, with the largest beta coefficient.

**Table 5.** Beta coefficients, standard errors, standardized betas and significance value for creatinine, phosphorus and uric acid as predictors of self-perceived cognitive state.

|  | **B** | **SE B** | **beta** | **P** |
| --- | --- | --- | --- | --- |
| Creatinine | -.337 | .350 | -.125 | .339 |
| Phosphorus | -.610 | .572 | -.135 | .290 |
| Uric Acid | -.781 | .458 | -.209 | .092 |

Abbreviations: B., Beta coefficients; SE., Standard errors.

#### 3.3.4. SELF-PERCEIVED PSYCHOLOGICAL WELL-BEING AND SOCIODEMOGRAPHIC VARIABLES

A one-way between-groups analysis of variance was conducted to examine the impact of sociodemographic variables on self-perceived psychological well-being. The results revealed a significant difference in the total psychological well-being score [*F*(1, 70) = 7.235, *p* = .009], with a worse self-perceived psychological well-being state in single patients (*M* = 33.08, *SD* = 8.95) compared to those who were in a relationship (*M* = 38.61, *SD* = 8.48). The effect size, calculated using eta squared, was .09. No other significant results were found for the remaining sociodemographic variables.

#### 3.3.5. SELF-PERCEIVED PSYCHOLOGICAL WELL-BEING AND CLINICAL VARIABLES

A one-way between-groups analysis of variance was conducted to explore the impact of clinical variables on self-perceived psychological state. Results did not reveal a significant difference in the total psychological well-being score. No significant results were identified.

#### 3.3.6. SELF-PERCEIVED PSYCHOLOGICAL WELL-BEING AND BIOCHEMICAL VARIABLES

The relationship between the selected biomarkers (creatinine, albumin, uric acid, phosphorus, C-reactive protein, hemoglobin, hemoglobin A1c, calcium, sodium, potassium) and the total score of self-perceived psychological well-being was examined using Pearson correlation coefficient. No significant correlations were found between these variables.

#### 3.3.7. OTHER CORRELATIONS

This section describes other relevant associations between variables, taking into account the objectives of the current study. Results from the exploratory analysis using the Pearson correlation coefficient are shown in Table 6. A strong positive correlation was observed between total well-being and total cognition scores (*r* = .523, *p* < 0.01), suggesting that higher perceived well-being is associated with better self-perceived cognitive state.

**Table 6.** Pearson correlation coefficients between clinical, biochemical, cognitive and psychological variables.

|  | BMI | Cholesterol<br>HDL | Hemoglobin | Creatinine | Phosphorus | Uric<br>Acid | Albumin | Calcium | C-Reactive<br>Protein | Cholesterol<br>Total | PTH | Triglycerides | Leukocytes | Total<br>Well-being<br>score | Total<br>Cognition<br>score |
| --- | --- | --- | --- | --- | --- | --- | --- | --- | --- | --- | --- | --- | --- | --- | --- |
| BMI | 1 | -.255*<br>.028 | .202<br>.076 | -.130<br>.258 | -.191<br>.094 | -.145<br>.211 | -.068<br>.553 | -.157<br>.169 | -.064<br>.585 | -.213<br>.066 | -.010<br>.931 | .180<br>.123 | .089<br>.436 | -.174<br>.128 | .095<br>.407 |
| Cholesterol<br>HDL |  | 1 | .003<br>.981 | -.035<br>.764 | .209<br>.072 | .126<br>.284 | .209<br>.072 | .012<br>.917 | -.274*<br>.018 | .513**<br>.000 | .168<br>.162 | -.310**<br>.007 | -.286<br>.013 | .019<br>.871 | -.165<br>.158 |
| Hemoglobin |  |  | 1 | .008<br>.945 | .056<br>.624 | .064<br>.586 | -.031<br>.790 | .017<br>.883 | -.111<br>.342 | .108<br>.354 | -.090<br>.444 | .272*<br>.018 | .093<br>.420 | -.043<br>.711 | -.153<br>.181 |
| Creatinine |  |  |  | 1 | .464**<br>.000 | .408**<br>.000 | .306<br>.007 | -.148<br>.197 | -.046<br>.697 | .201<br>.083 | .180<br>.124 | .054<br>.644 | -.038<br>.744 | -.082<br>.476 | -.250*<br>.027 |
| Phosphorus |  |  |  |  | 1 | .341**<br>.003 | .098<br>.393 | -.131<br>.254 | .022<br>.849 | .269*<br>.019 | .257*<br>.027 | .035<br>.766 | .097<br>.396 | -.105<br>.359 | -.276*<br>.014 |
| Uric Acid |  |  |  |  |  | 1 | .187<br>.105 | -.228<br>.048 | .030<br>.802 | .114<br>.333 | .048<br>.687 | -.037<br>.755 | -.049<br>.675 | -.004<br>.974 | -.306**<br>.007 |
| Albumin |  |  |  |  |  |  | 1 | .357**<br>.001 | -.283*<br>.014 | .255*<br>.027 | .008<br>.944 | .101<br>.388 | -.095<br>.408 | -.194<br>.090 | -.176<br>.122 |
| Calcium |  |  |  |  |  |  |  | 1 | -.077<br>.511 | .135<br>.249 | -.013<br>.911 | -.045<br>.704 | -.031<br>.788 | .006<br>.959 | .128<br>.264 |
| C-Reactive<br>Protein |  |  |  |  |  |  |  |  | 1 | -.045<br>.706 | -.196<br>.100 | .061<br>.607 | .185<br>.113 | .190<br>.103 | .201<br>.084 |
| Cholesterol<br>Total |  |  |  |  |  |  |  |  |  | 1 | .237*<br>.046 | .138<br>.239 | -.128<br>.273 | .020<br>.868 | -.111<br>.344 |
| PTH |  |  |  |  |  |  |  |  |  |  | 1 | .020<br>.866 | -.084<br>.478 | -.014<br>.908 | -.075<br>.524 |
| Triglycerides |  |  |  |  |  |  |  |  |  |  |  | 1 | .378**<br>.001 | -.140<br>.232 | .007<br>.950 |
| Leukocytes |  |  |  |  |  |  |  |  |  |  |  |  | 1 | .049<br>.670 | .167<br>.143 |
| Total Well-being score |  |  |  |  |  |  |  |  |  |  |  |  |  | 1 | .523**<br>.000 |
| Total<br>Cognition<br>score |  |  |  |  |  |  |  |  |  |  |  |  |  |  | 1 |

\*\* $p < 0.01$ / \*
$p < 0.05$ . Abbreviations: HDL., High-density lipoprotein; PTH., Parathyroid hormone.

## 4. DISCUSSION

This study aimed to assess the self-perceived cognitive and psychological well-being of individuals undergoing HD and to explore its association with sociodemographic, clinical, and biochemical variables.

Our results showed that higher levels of creatinine, phosphorus, and uric acid were significantly associated with poorer cognitive self-perception. Relationship status and the presence of diabetes were also linked to subjective cognition. However, psychological well-being was not significantly correlated with biochemical markers, being instead influenced by social factors, such as relationship status. These findings highlight the multifactorial nature of cognitive and emotional symptoms in CKD, highlighting the importance of a holistic approach to patient care.

In our study, a very high percentage of the participants (89.7%) had hypertension, with more than half of these (51.3%) also suffering from diabetes. This demonstrates the increasing prevalence of CKD in developing countries driven by the significant increase in lifestyle-related non-infectious diseases, such as diabetes and hypertension. These findings are consistent with previous research indicating that diabetes and hypertension are the leading causes of CKD in developed countries (Jha et al., 2013).

### 4.1. SELF-PERCEIVED COGNITIVE AND PSYCHOLOGICAL WELL-BEING

As previously observed, the literature consistently indicates that a significant proportion of individuals undergoing HD (estimated to be between 70% and 80%) experience cognitive impairment (Andrews et al., 2025), and mood disorders such as anxiety and depression are frequently observed (Sharma et al., 2022). Nevertheless, participants in this study self-reported relatively high levels of both cognitive and psychological well-being. This apparent discrepancy does not necessarily contradict previous findings; however, it reflects the difference between objective clinical assessments and subjective self-perception. While this discrepancy between patients’ perceptions and clinical findings highlights the limitations of relying solely on self-reports, it can be explained in light of several contributing factors.

Firstly, subjective cognitive complaints (SCCs) do not always align with objective performance-based measures. As Chan et al. (2022) noted, SCCs in patients on HD tend to be more closely associated with emotional symptoms such as anxiety and depression than with actual cognitive test results. Patients who feel emotionally stable or well-supported may perceive their cognitive state positively, even when some impairment is present. Secondly, common coping mechanisms such as adaptation, minimization, or optimistic coping, may lead patients to overlook psychological or cognitive difficulties (Mesa-Gresa et al., 2023; Thongon et al., 2022). Interestingly, our findings are consistent with those of Mesa-Gresa et al. (2023) who also found that patients and their carers had positive perceptions of the patients’ cognitive and psychological state, even though other aspects of their study revealed underlying feelings of frustration, fatigue, and emotional strain. Unlike our study, which focused exclusively on 78 patients’ self-reports, enabling us to explore specific associations between psychological and cognitive variables and clinical indicators, Mesa-Gresa et al. (2023) also included perspectives from carers and healthcare professionals, providing a different perspective on the patients’ conditions. This broader approach highlighted discrepancies between how patients perceive themselves and how others perceive their well-being, underscoring the importance of interpreting subjective reports within a wider psychosocial and clinical framework (Elias et al., 2025).

Thirdly, qualitative evidence suggests that the structured nature of dialysis may provide a temporary sense of stability, masking functional limitations and psychological distress through adaptation and cognitive reframing (Chan et al., 2025; Elias et al., 2025).

Future studies should complement subjective self-assessments with objective neuropsychological testing to provide a more accurate evaluation of cognitive status in individuals with CKD 5D.

### 4.2. ASSOCIATION OF SELF-PERCEIVED COGNITIVE STATE WITH SOCIODEMOGRAPHIC, CLINICAL, AND BIOCHEMICAL VARIABLES

Our findings are consistent with previous studies indicating that cognitive function is frequently impaired in individuals with CKD undergoing HD (Giannakou et al., 2023). In our sample, being single and not having diabetes were both associated with poorer self-perceived cognitive functioning. The fact that in our study single patients reported poorer cognitive status may reflect reduced psychosocial support, a factor known to influence cognitive health. Silberman-Beltramella et al. (2022) observed that older Spanish adults living with a partner had better physical, functional, and cognitive health scores, though the latter was not statistically significant. Their study highlighted loneliness as the strongest predictor of poor health, suggesting that partnership may indirectly protect cognition by buffering against social isolation and reducing perceived loneliness, both of which are key factors in healthy aging (Miao et al., 2025). Taken together, these findings imply that social connection and partnership may indirectly protect both mental and cognitive well-being in CKD 5D by mitigating loneliness and providing social support (Sułkowski et al., 2024).

Previous research has shown that individuals with diabetes tend to report more cognitive complaints (Chan et al., 2022), and cognitive impairment related to diabetes is now widely recognized as a major complication, with substantial impact on patients’ overall HRQoL (Yu et al., 2025). Interestingly, in our sample, the absence of diabetes was associated with poorer self-perceived cognitive functioning. While diabetes is a well-established risk factor for cognitive decline in CKD 5D (Joseph et al., 2019), this discrepancy could be explained by the differences in health perception, symptom attribution biases or illness perception. The “biographical disruption” associated with dialysis which is marked by unexpected illness, altered identity, and emotional distress followed by adaptation and cognitive reframing (Elias et al., 2025), may explain greater cognitive concern among non-diabetic patients. Without prior framing, cognitive changes may appear unfamiliar or threatening, whereas diabetic patients may interpret similar difficulties as part of an expected disease burden or ageing.

Significant negative correlations were observed between cognitive self-assessment and three key biomarkers: creatinine, phosphorus, and uric acid. Previous studies have linked elevated creatinine levels in CKD 5D to structural changes in brain regions such as the insula and temporal lobe, which are crucial for cognitive functioning (Fang et al., 2024). Similarly, since phosphorus is essential for energy metabolism and brain health, both insufficient and excessive phosphorus levels are associated with cognitive deficits. Insufficient phosphorus levels are linked to an increase in cerebral β-amyloid accumulation, a characteristic feature of Alzheimer’s disease (Park et al., 2017), while the latter may contribute to the onset of dementia (Li et al., 2017). Uric acid, a molecule with both antioxidant and pro-inflammatory properties, also showed a negative association with cognitive self-assessment. Although some studies highlight the neuroprotective role of this molecule (Ye et al., 2016), it has also been linked to increased inflammation and cognitive dysfunction in CKD 5D (Mijailovic et al., 2022). The regression analysis performed with our data further supported this relationship, as uric acid contributed the most to the model predicting cognitive self-perceived state, though none of the predictors reached individual statistical significance. The model explained 9.8% of the variance in self-perceived cognitive function, indicating a modest yet meaningful association between these biochemical markers and cognitive outcomes. This supports a multifactorial view of cognitive symptoms in CKD and underscores the value of multimodal assessment that integrates biochemical indicators with patient-reported outcomes (Wang et al., 2023).

### 4.3. ASSOCIATION OF SELF-PERCEIVED PSYCHOLOGICAL WELL-BEING WITH SOCIODEMOGRAPHIC, CLINICAL, AND BIOCHEMICAL VARIABLES

The significant positive correlation observed between total well-being and total cognition underscores the close interrelationship between emotional and cognitive functioning in individuals with CKD 5D. This finding suggests that patients who perceive themselves as feeling better emotionally may also report greater cognitive functioning, which aligns with existing literature on the bidirectional influence between mood and cognition in chronic illness. Emotional intelligence also plays a critical role, as it integrates emotional regulation with cognitive functioning and is consistently linked to higher levels of psychological well-being (Gutiérrez-Cobo et al., 2016). Moreover, concerns about cognitive decline, such as fear of Alzheimer’s disease, are associated with reduced emotional well-being (Cutler & Brăgaru, 2017) and emotional disorders like anxiety and depression can impair cognitive control, particularly in affective regulation (Chau et al., 2025).

In the context of CKD 5D, these interconnections become particularly relevant, as the disease’s impact on both psychological well-being and cognitive health is shaped not only by internal traits but also by social and clinical factors. The high incidence of anxiety, depression, and psychological distress in people undergoing HD highlights the significant impact of the disease on mental well-being, in addition to its physiological adversities. In this regard, Yoong et al. (2017) reported a higher risk of clinical depression in patients who were single, suggesting that sociocultural factors may help identify patients at risk of depression. In the present study, unlike cognitive outcomes, psychological well-being was not significantly correlated with biochemical parameters. Instead, it was influenced by sociodemographic factors, such as relationship status. This finding is consistent with previous research suggesting that emotional distress is often shaped by social context and disease characteristics (Bossola et al., 2010; Yoong et al., 2017). Similarly, marital status can act as a protective factor for HRQoL in CKD patients (Gerogianni & Babatsikou, 2014), likely due to the buffering role of emotional support provided by intimate relationships. These findings emphasize the importance of considering relationship status and patient history when evaluating emotional health in CKD 5D.

### 4.4. STRENGTH AND LIMITATIONS

One of the strengths of this study lies in its focus on patients’ subjective perceptions of their cognitive and emotional health, an area often overlooked in nephrology research (Chan et al., 2022; Chan et al., 2024). The use of ad-hoc, professionally developed questionnaires tailored for patients on HD adds contextual relevance to the findings (Mesa-Gresa et al., 2023). However, several limitations must be acknowledged. First, assessments were conducted during dialysis sessions, which may have introduced noise and stress-related bias. Second, the cross-sectional design limits causal interpretations. Third, there was a lack of objective neuropsychological testing, which restricts comparison with clinical diagnostic criteria for cognitive impairment.

### 4.5. RECOMMENDATIONS FOR FUTURE RESEARCH

This study underscores the importance of integrating subjective assessments into routine care for patients undergoing HD. Given the modest but significant influence of certain biomarkers on cognitive perception, future research should consider combining subjective, biochemical, and objective neurocognitive measures. Longitudinal designs with larger samples and standardized cognitive testing could help clarify the causal pathways linking CKD, metabolism, and mental health. Finally, knowing that physical activity or psychosocial interventions have been identified as valuable tools to enhance HRQoL in this population (Marin et al., 2023), future studies should aim to improve patient outcomes in CKD not by medical management, but also by implementing interventions that address emotional well-being and cognitive health.

## 5. CONCLUSIONS

This study highlights the complex and multifactorial nature of cognitive functioning and psychological well-being in individuals undergoing HD. While participants reported relatively high self-perceived cognitive and emotional health, this contrasts with existing clinical evidence suggesting high prevalence of cognitive impairment and mood disorders in this population. The findings underscore the need to interpret subjective assessments within a broader biopsychosocial context, accounting for emotional support, treatment environment, and possible effects of polypharmacy.

Self-perceived cognitive state was associated with biochemical indicators of kidney dysfunction (creatinine, phosphorus, and uric acid), as well as sociodemographic (relationship status) and clinical (the presence of diabetes) factors. In contrast, psychological well-being was shaped more by social characteristics, rather than by biochemical markers. These patterns reinforce the importance of integrating both objective clinical indicators and patient-reported outcomes in the assessment of health status.

Altogether, the findings support a multidimensional model of care that combines medical monitoring with psychosocial assessment and intervention. Efforts to improve outcomes in CKD 5D should include strategies to protect and enhance patients’ cognitive and emotional health as well as managing the disease.

## Author contributions

Conceptualization, P.M.-G., R.R., and A.-E.M.; methodology, A.-E.M.; data collection, A.-E.M. and A.G.-T.; validation, A.-E.M. and A.G.-T.; formal analysis, A.-E.M. and P.M.-G.; investigation, A.-E.M.; resources, A.-E.M. and E.S.-O.; data curation, A.-E.M.; writing—original draft preparation, A.-E.M.; writing—review and editing, P.M.-G., R.R., A.G.-T. and E.S.-O.; visualization, A.-E.M.; supervision, P.M.-G.; project administration, P.M.-G. All authors have read and agreed to the published version of the manuscript.

## Conflict of Interest

The authors declare no conflict of interest.

## Data availability

The data that support the findings of this study are available from the corresponding author upon reasonable request.

## Funding statement

Research reported in this publication was supported by MCIN/AEI/10.13039/501100011033/ and by “ERDF A way of making Europe” under grant number PID2022-138021OA-I00, as well as the GoodRENal project, which is funded by the European Community Erasmus+ (SRIC_UE_2020-1-ES01-KA204-083141).

## Notes

### Competing Interest Statement

The authors have declared no competing interest.

### Author Declarations

The study adhered to the principles outlined in the Declaration of Helsinki and was approved by the Ethics Committee of the University of Valencia (Reference: 2023-PSILOG-2558999) and the Hospital Universitario y Politecnico La Fe (Spain) (Registration number: 2020-064-1).

